# A systematic review on diagnostic and prognostic biomarkers for bladder cancer

**DOI:** 10.1101/2024.06.02.24308331

**Authors:** Umar Muhammad, Umar Ahmad, Buhari Ibrahim, Aliyu Adamu Ahmad, Haruna Usman Liman

## Abstract

**Background:** Bladder cancer is one of the most prevalent malignancies worldwide. Despite its high incidence, public awareness of the condition remains low, and it has received less research attention compared to other common cancers. Over the past 80 years, patient outcomes and treatment strategies have remained largely unchanged, with cystoscopy being the primary method for detecting bladder cancer. This procedure, often repeated during long-term surveillance due to the recurrent nature of bladder tumors, is both uncomfortable for patients and costly for healthcare providers. The identification and validation of molecular biomarkers in blood, urine, or tissue could facilitate tumour detection and reduce reliance on cystoscopy.

**Aim:** This study aims to identify potential molecular biomarkers for bladder cancer that could improve tumour detection and lessen the need for repeated cystoscopies.

**Methods:** A systematic review was conducted, searching for articles related to bladder cancer biomarkers in four databases: PubMed, ScienceDirect, Google Scholar, and Cochrane. Studies that met the inclusion criteria underwent title/abstract screening and full-text review. A total of twenty studies were deemed eligible for inclusion in this review.

**Results:** The review identified several gene product biomarkers, including TEAD4, TPM1, TPM2, SKA3, EO1, HYAL3, MTDH, EPDR1, hTERT, KRT7, SW, ARHGAP9, XPH4, OTX1, BUB1, and Usp28. Additionally, protein product biomarkers were identified, such as A1AT, APOE, AG, CA9, IL8, MMP9, MMP10, PAI1, SCDI1, SDC1, VEGFA, CD73, TIP2, CXCL5, PCAT6, and NCR3LG1 (B7-H6).

**Conclusion:** The study highlights the potential of various gene and protein biomarkers for the detection of bladder cancer. Further research is necessary to validate these biomarkers’ diagnostic and prognostic potential in identifying bladder cancer in suspected cases.

## 1.0 Introduction

Bladder cancer ranks as the tenth most prevalent cancer globally, disproportionately affecting males four times more frequently than females (Lobo *et al*., 2022). In 2020, approximately 573,000 cases of bladder cancer were reported, according to Global Cancer Incidence, Mortality, and Prevalence (GLOBOCAN) data (Tataroğlu, 2022). Tobacco use is the primary recognized cause, responsible for up to two-thirds of all bladder malignancies and 30–40% of urothelial carcinoma cases (Tataroğlu, 2022). With the world’s population projected to exceed 9.7 billion by 2050, the incidence rate of bladder cancer is expected to rise, placing a significant burden on healthcare systems worldwide (Kourie *et al*., 2022).

Bladder cancer is predominantly induced by cigarette smoking, advanced age, and male sex. The disease can present with microscopic hematuria, detected through cystoscopy and upper urinary tract imaging, which remain the gold standard for initial diagnosis (Lenis *et al*., 2020). Genetic factors also play a role, with mutations in tumor suppressor genes such as PTEN and MSH2, linked to Cowden and Lynch syndromes, increasing the incidence of urothelial and squamous bladder cancer (Tataroğlu, 2022).

Treatment for non-muscle-invasive bladder cancer (NMIBC) involves endoscopic surgery and therapeutic interventions tailored to the risk of malignancy (de Ruiter *et al*., 2022). Advanced technology in cystoscopy improves tumor detection and reduces recurrence risk (IBINGIRA *et al*., 2022). For muscle-invasive bladder cancer (MIBC), more aggressive treatments such as radical surgery, urine diversion, or trimodal therapy involving surgery, chemotherapy, and radiation are recommended (de Ruiter *et al*., 2022). Chemotherapy options continue to evolve, providing alternatives for patients with varying disease stages (Ertl *et al*., 2022).

Recent advancements in diagnostic techniques include ultrasonography, intravascular urography (IVU), computed tomography (CT), magnetic resonance imaging (MRI), biopsy, and cytology (Konala *et al*., 2022). Fluorescence-controlled diagnostics offer exceptional precision and sensitivity (Compérat *et al*., 2022). Despite these advancements, the translation of genetic findings, particularly for NMIBC, into clinical practice remains limited (Malinaric *et al*., 2022).

The emergence of precision medicine aims to address this gap by considering patient and tumor diversity to enable targeted therapies using potential biomarkers (Shimura *et al*., 2021). Biomarkers hold the promise of improving diagnostic accuracy, predicting treatment outcomes, and enhancing therapeutic responses (Vlachostergios & Faltas, 2019). Numerous biomarkers have been investigated for their roles in screening, surveillance, and follow-up (Yutkin *et al*., 2010). However, despite significant efforts, no biomarkers have yet been universally approved and applied in clinical practice (Yutkin *et al*., 2010). The accurate application of potential biomarkers hinges on their reliability and validity (Grossman *et al*., 2019).

This systematic review aims to identify and evaluate potential diagnostic and prognostic biomarkers for bladder cancer. By consolidating findings from recent studies, this review seeks to advance the understanding of biomarkers that could significantly impact the clinical management of bladder cancer.

## 2.0 Materials and methods

### 2.1 Study design

This review was designed based on the Population, Intervention, Comparisons, and Outcomes (PICOS) framework. The population comprised patients with the presence of bladder cancer. The interventions involved the identification and analysis of biomarkers. The comparator was either diagnostic or prognostic methods. The outcomes focused on the presence or absence of potential biomarkers that could determine the presence of bladder cancer.

### 2.2 Protocol and registration

The study was registered with PROSPERO, and the registration ID is pending. The Preferred Reporting Items for Systematic Reviews and Meta-Analyses (PRISMA) guidelines were followed to ensure a systematic and transparent review process.

### 2.3 Literature search strategy

A systematic review of the literature was conducted in accordance with Cochrane Methods for systematic reviews(Henderson *et al.,* 2010). The databases PubMed (White, 2020), ScienceDirect (Gies, 2018), Google Scholar (Halevi *et al*., 2017), and Cochrane (Kleinstäuber *et al*., 1996), were searched for articles published between January 2012 and November 2022.

Two reviewers, Umar Muhammad and Aliyu Adamu Ahmad, independently conducted the literature search using the following keyword strategy: (((diagnostic) OR (prognostic)) OR (biomarkers)) OR (bladder cancer). Filters were applied to restrict results to the period from 2012 to 2023. Specific search terms included:

A. “diagnosis” [MeSH Terms] OR “diagnosis” [All Fields] OR “diagnostic” [All Fields] OR “diagnostical” [All Fields] OR “diagnostically” [All Fields] OR “diagnostics” [All Fields]
B. “prognostic” [All Fields] OR “prognostical” [All Fields] OR “prognostically” [All Fields] OR “prognosticate” [All Fields] OR “prognosticated” [All Fields] OR “prognosticates” [All Fields] OR “prognosticating” [All Fields] OR “prognostication” [All Fields] OR “prognostications” [All Fields] OR “prognosticator” [All Fields] OR “prognosticators” [All Fields] OR “prognostics” [All Fields]
C. “biomarkers” [All Fields] OR “biomarkers” [MeSH Terms] OR “biomarkers” [All Fields] OR “biomarker” [All Fields]
D. “urinary bladder neoplasms” [MeSH Terms] OR (“urinary” [All Fields] AND “bladder” [All Fields] AND “neoplasms” [All Fields]) OR “urinary bladder neoplasms” [All Fields] OR (“bladder” [All Fields] AND “cancer” [All Fields]) OR “bladder cancer” [All Fields]

The search results were exported and managed using the online systematic review software Covidence (Babineau, 2014). Duplicate articles were removed, and the remaining articles underwent further screening based on titles and abstracts. Full-text articles were then reviewed to identify studies that met the inclusion criteria.

### 2.4 Study selection

The study selection process was managed using the online systematic review software Covidence (https://www.covidence.org)(Babineau, 2014). This tool facilitated the organization and screening of articles retrieved from the bibliographic databases. The articles were independently screened by two reviewers based on their titles, abstracts, and the availability of full-texts.

#### 2.4.1 Inclusion criteria

A. Original research articles focused on bladder cancer.
B. Published in English.
C. Various study designs and participant demographics.
D. Studies investigating genes or proteins in blood, urine, or tissue samples.
E. Comparison of the diagnostic or prognostic value of these biomarkers, including cystoscopy as a comparator.
F. Desired outcomes included the presence or absence of potential biomarkers and their accuracy.

#### 2.4.1 Exclusion criteria

A. Non-original research articles.
B. Studies unrelated to bladder cancer biomarkers.
C. Studies lacking clarity on identified biomarkers.
D. Studies without a comparison group.
E. Animal or in vitro studies.
F. Articles not published in English.

The specified publication date range was between January 2012 and November 2022. This rigorous screening process ensured the selection of high-quality studies relevant to the systematic review’s objectives, ultimately including those that met all predefined criteria.

### 2.5 Data extraction

This systematic review strictly followed the PRISMA guidelines during the data extraction phase to ensure a transparent and systematic process. Two independent reviewers meticulously gathered information on key aspects of the studies, including study design, participant characteristics, investigated biomarkers, outcomes, and statistical methods. Any discrepancies in the extracted data were resolved through consensus or, if necessary, by consulting a third reviewer, thereby maintaining the integrity and accuracy of the process. The initial data extraction was carried out by the first reviewer, who carefully collected data from the included studies. This information was then cross-checked and confirmed by the second reviewer to ensure accuracy and completeness. The extracted data was compiled into a comprehensive table, structured according to PRISMA standards.

### 3.6 Quality assessment

The quality of the included studies was independently evaluated by two reviewers using the Quality Assessment of Diagnostic Accuracy Studies (QUADAS-2) tool (Wade *et al*., 2013). This tool is specifically designed for assessing the risk of bias in observational studies, such as cohort and case-control studies (Wade *et al*., 2013). The QUADAS-2 tool evaluates four key domains:

1. **Patient Selection**: Examines the methods used to select patients and assesses whether these methods could introduce bias.
2. **Index Test**: Assesses the risk of bias in the conduct and interpretation of the test being evaluated.
3. **Reference Standard**: Evaluates the risk of bias associated with the application and interpretation of the reference standard.
4. **Flow and Timing**: Considers the potential for bias related to the timing of the index test and reference standard, as well as the flow of patients through the study.

Each domain is rated for risk of bias as “low,” “high,” or “some concerns.” A domain is judged to have a low risk of bias if all questions for that domain are answered affirmatively. If any questions are answered negatively, the risk of bias for that domain is considered high. The “some concerns” category is used when there is insufficient data to make a clear judgment.

The independent assessments by the two reviewers were compared, and any discrepancies were resolved through discussion or consultation with a third reviewer. This rigorous process ensured a thorough and unbiased evaluation of the included studies, enhancing the reliability and validity of the review’s conclusions.

## 3.0 Results

### 3.1 Literature search

A comprehensive literature search was conducted following the Preferred Reporting Items for Systematic Reviews and Meta-Analyses (PRISMA) guidelines. The objective was to identify relevant articles on bladder cancer biomarkers published from January 2012 to November 2022. The search encompassed four major databases: PubMed, ScienceDirect, Google Scholar, and Cochrane.

The search strategy employed medical subject headings (MeSH) and keywords related to bladder cancer, biomarkers, and diagnostic or prognostic outcomes. Terms included, but were not limited to, “bladder cancer,” “biomarkers,” “diagnostic,” “prognostic,” and their variations. Boolean operators “AND” and “OR” were used to refine the search results. Additionally, the reference lists of pertinent articles were hand-searched to uncover any studies that the initial electronic search might have missed.

Two independent reviewers, Umar Muhammad and Aliyu A. Ahmad, conducted the literature search to ensure comprehensive coverage and minimize bias. The search yielded a total of 2950 articles. Following this, approximately 650 duplicates were removed, leaving 2300 articles for further screening.

The remaining articles underwent a rigorous screening process based on their titles and abstracts, which resulted in the exclusion of a significant number of studies. The full texts of the remaining articles were then thoroughly reviewed. This process led to the exclusion of additional articles that did not meet the inclusion criteria. Ultimately, 35 articles were assessed for conformity with our inclusion criteria, of which 15 were excluded for various reasons.

Following this comprehensive screening process, 20 articles were deemed eligible and included in the final quantitative synthesis. These studies formed the basis for the systematic review and comprehensive analysis of bladder cancer biomarkers. The entire literature search process is summarized in Figure 1, which adheres to PRISMA standards, detailing the databases searched and the number of articles screened and selected for the systematic review.

**Figure 1:**
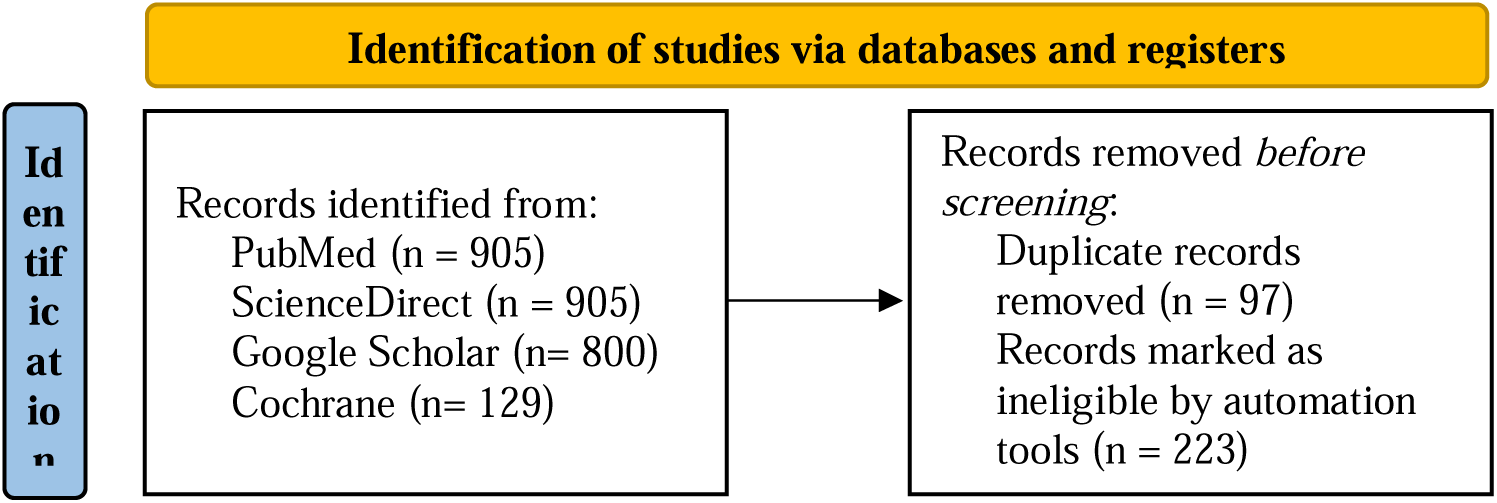
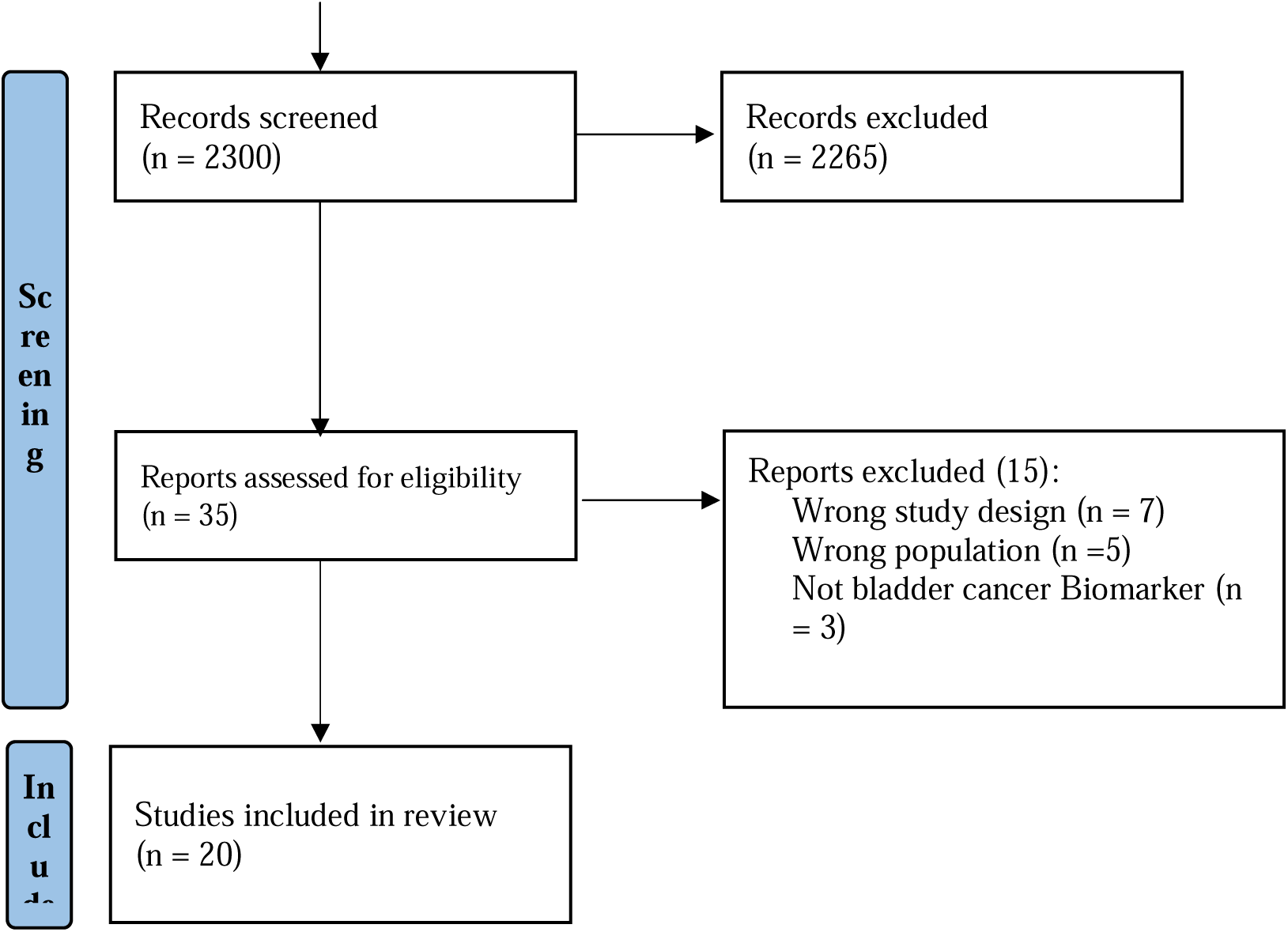
PRISMA flow diagram. PRISMA flow diagram illustrating the systematic review process, detailing the database searches and the number of articles screened, assessed for eligibility, and included in the qualitative synthesis. Out of 2950 articles identified through database searches, 20 articles met the inclusion criteria and were selected for qualitative analysis.

**Figure 2:**
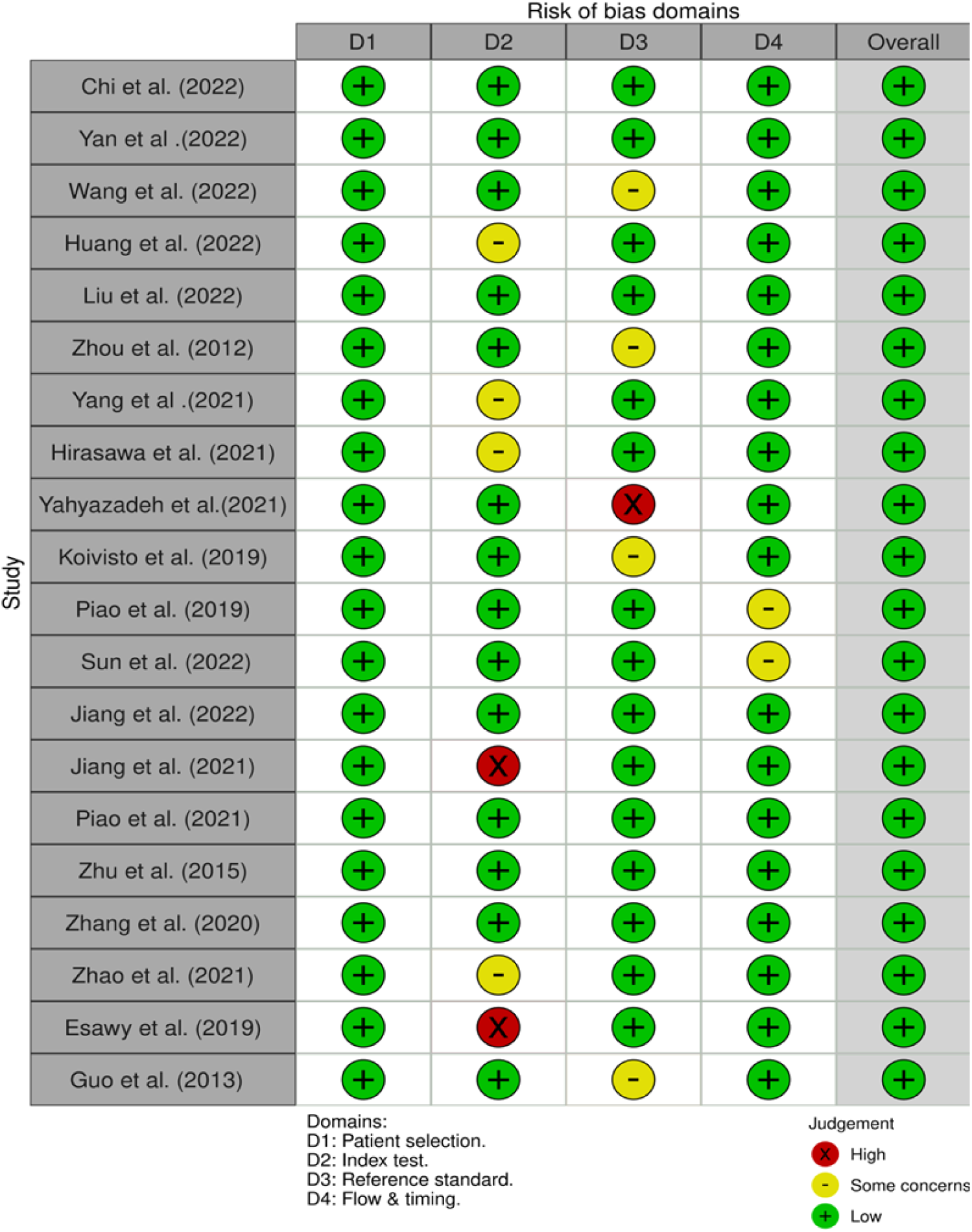
Evaluation of Bias Risk in Included Prospective Studies Using the Newcastle-Ottawa Scale.

### 3.2 Characteristics of the data extracted from the studies

The data extracted from the twenty selected studies focused on identifying diagnostic and prognostic biomarkers for bladder cancer, spanning from January 2012 to November 2022 (Table 1). These studies included original research articles with varied designs, such as cohort and case-control studies, conducted across multiple countries, including China, Korea, Iran, Finland, and the USA. This geographical diversity underscores the global interest in enhancing bladder cancer diagnosis and management through biomarker identification.

**Table 1:**
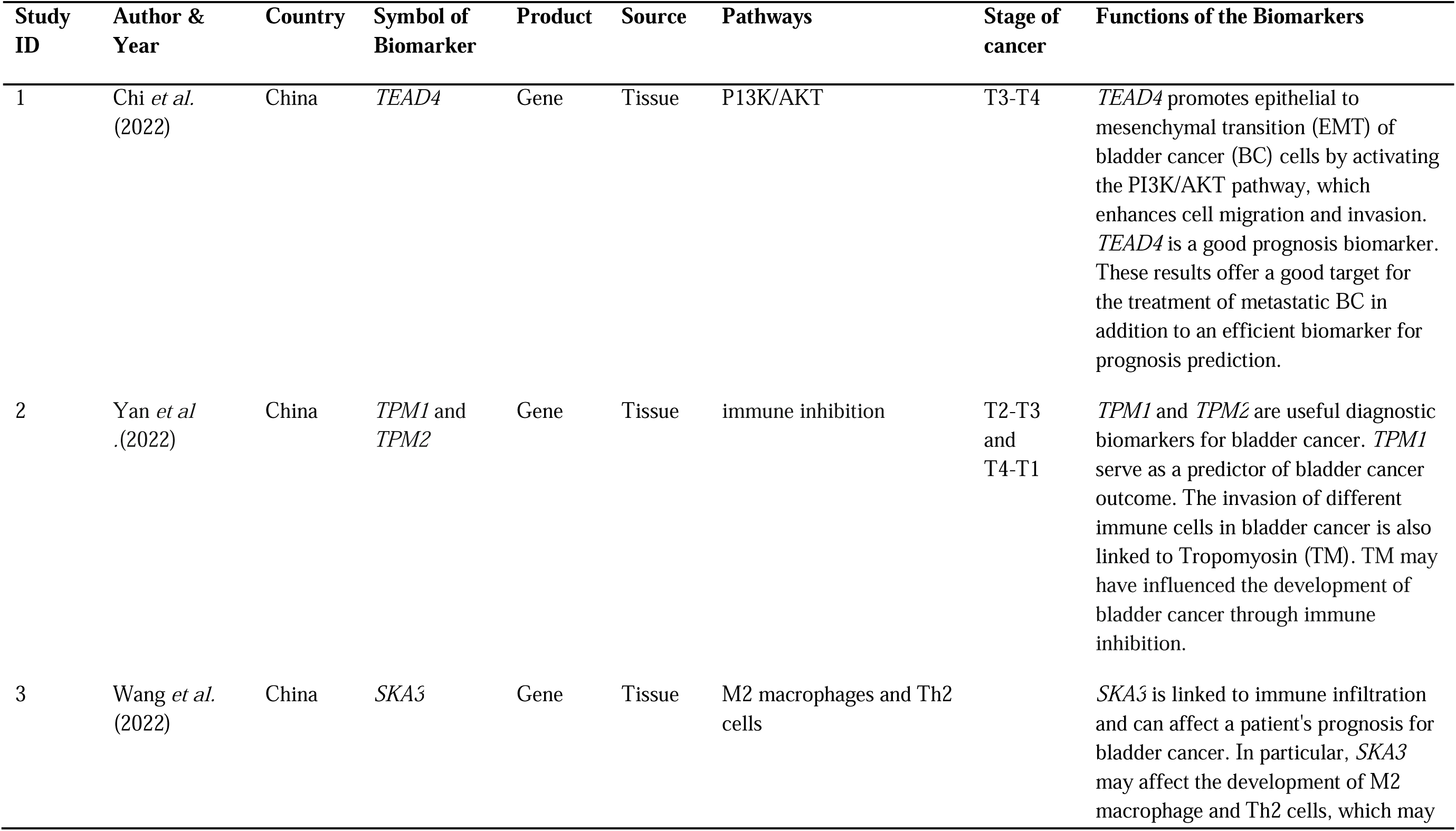
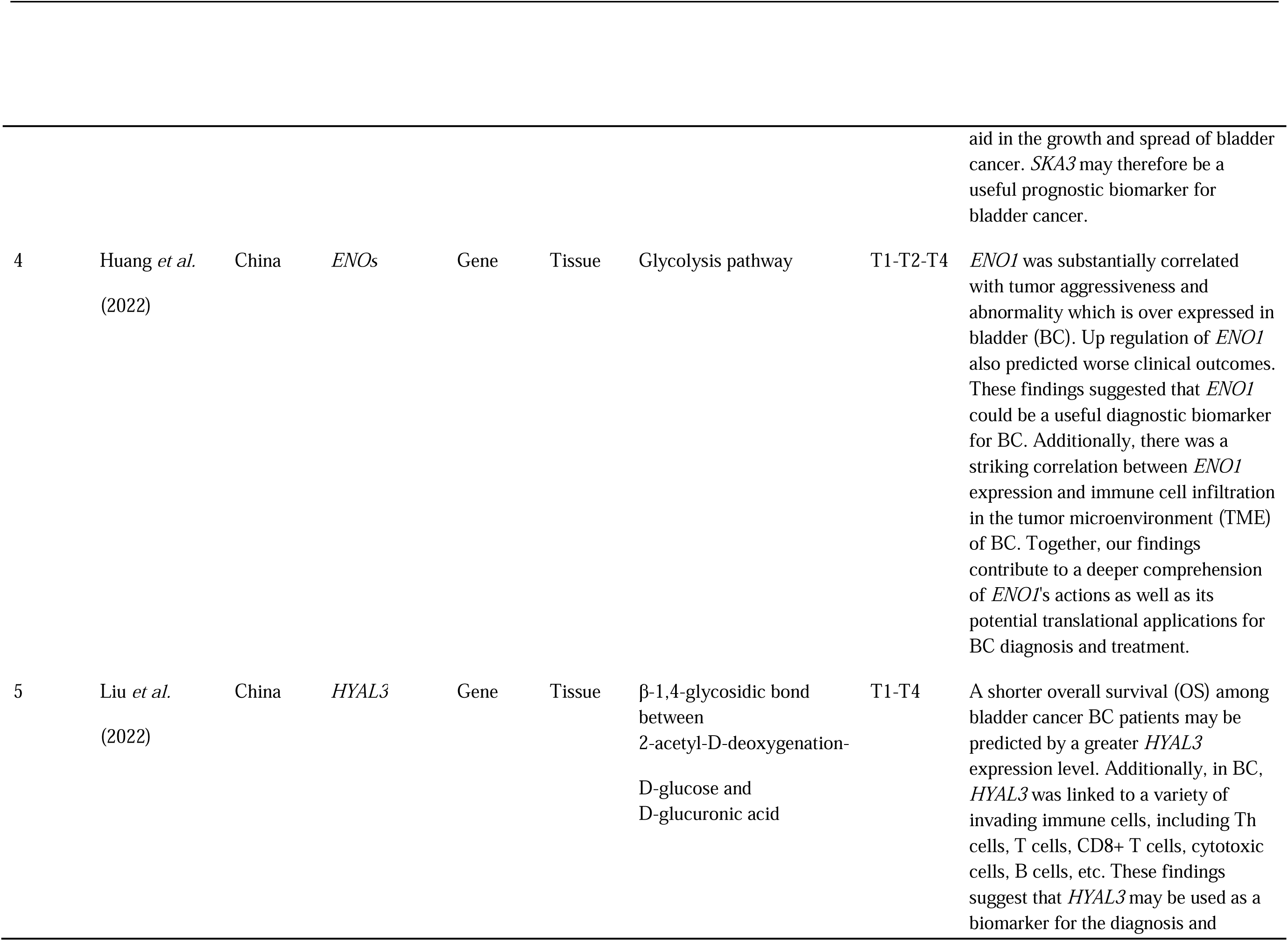
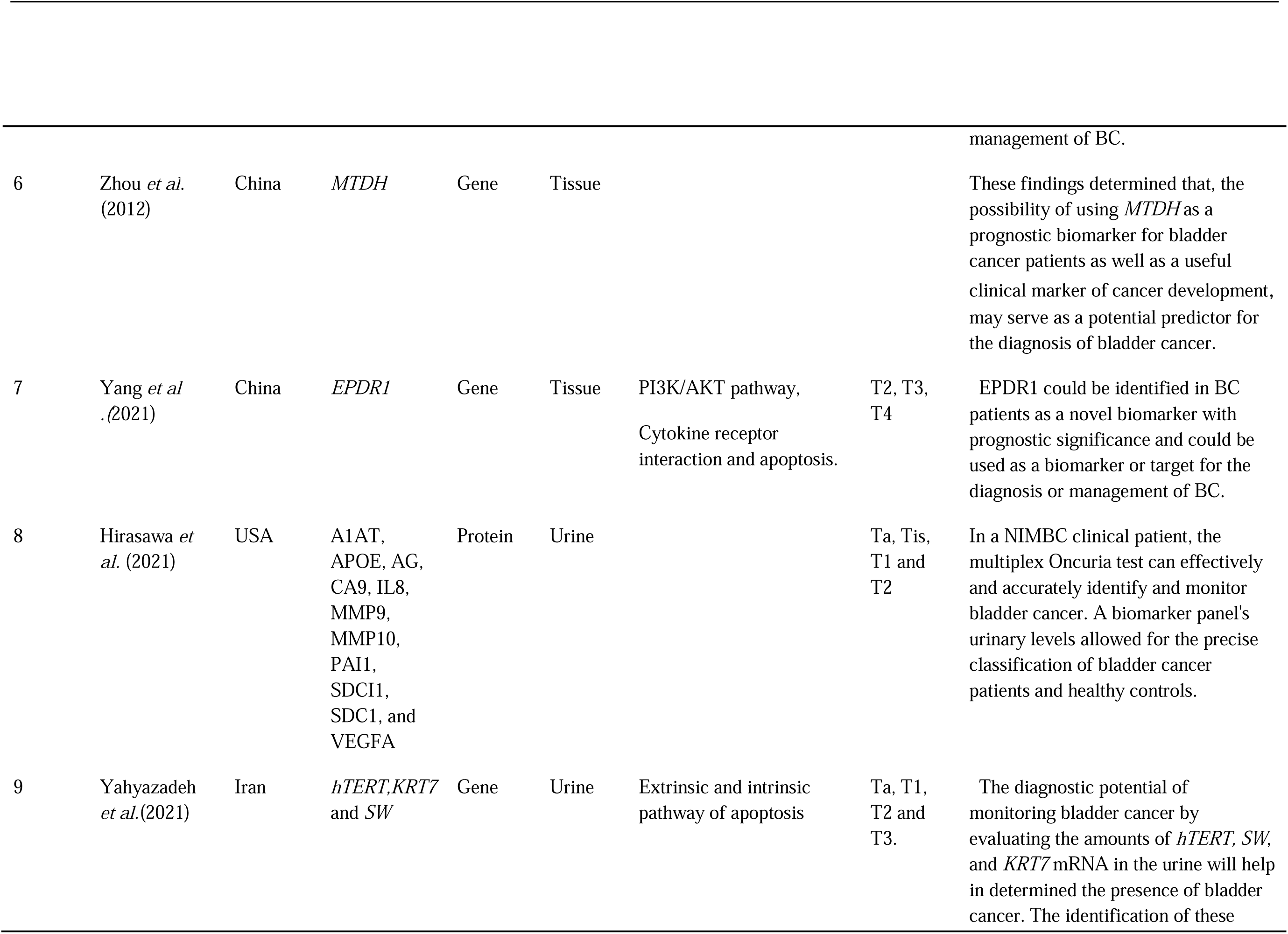
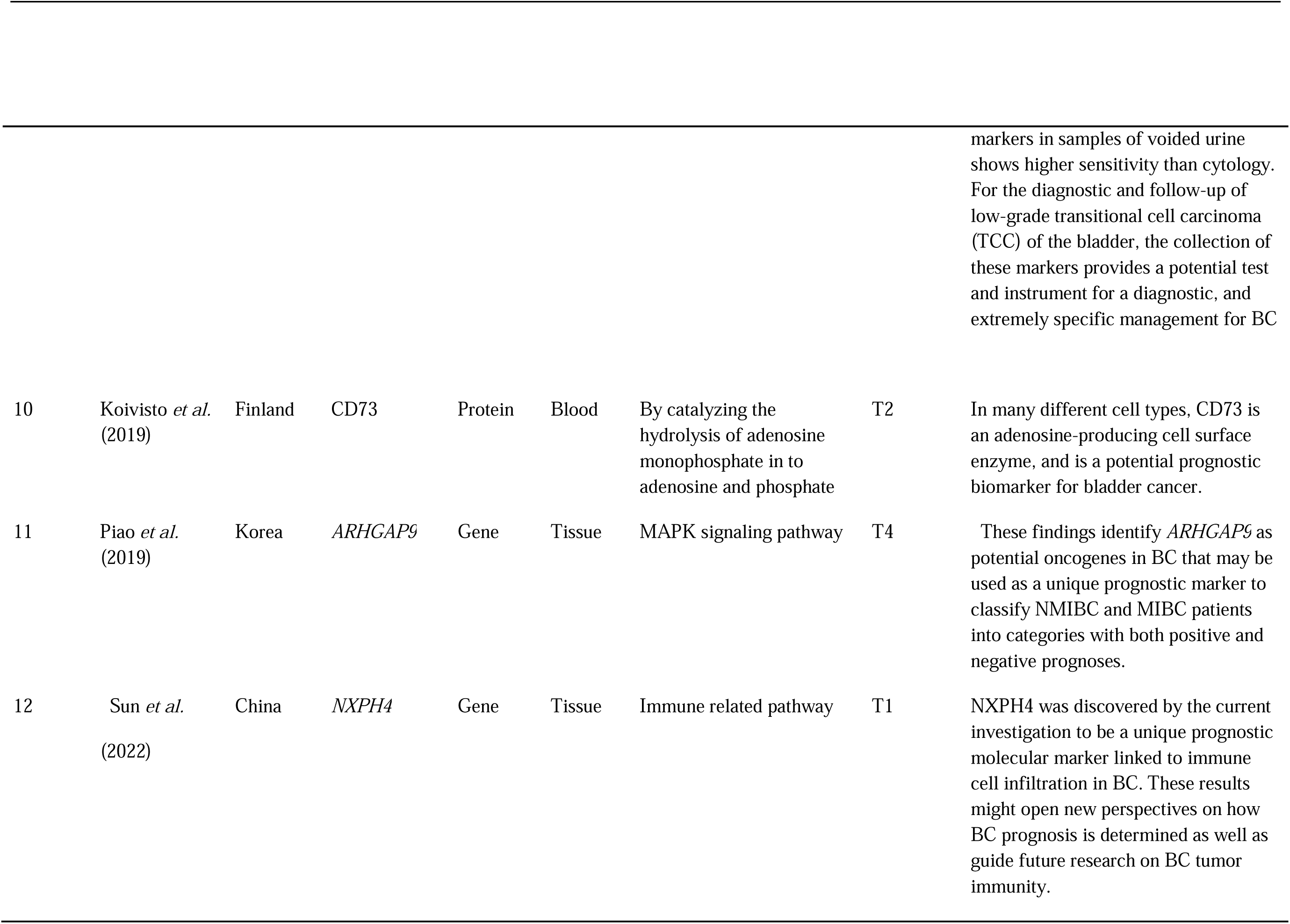
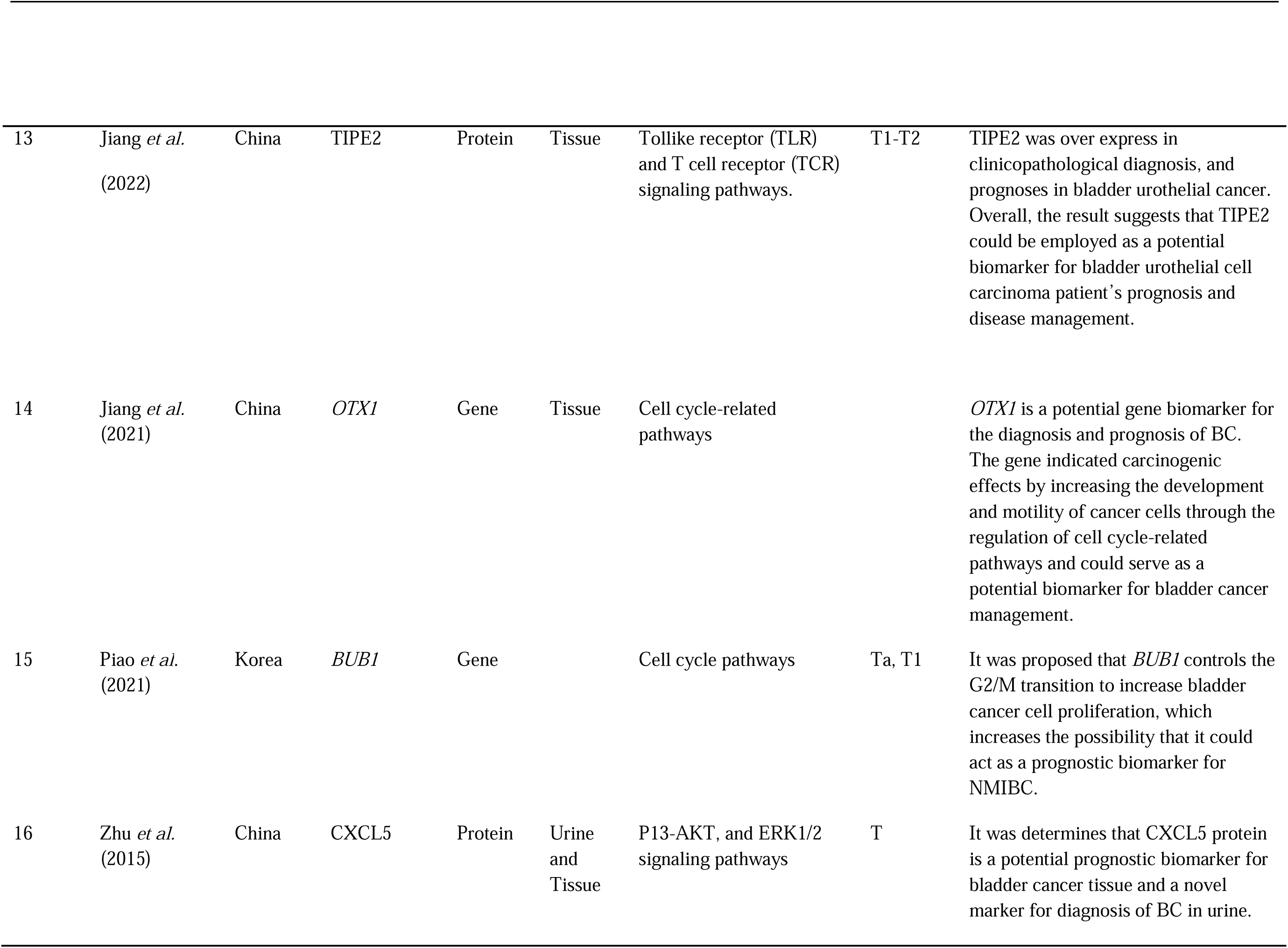
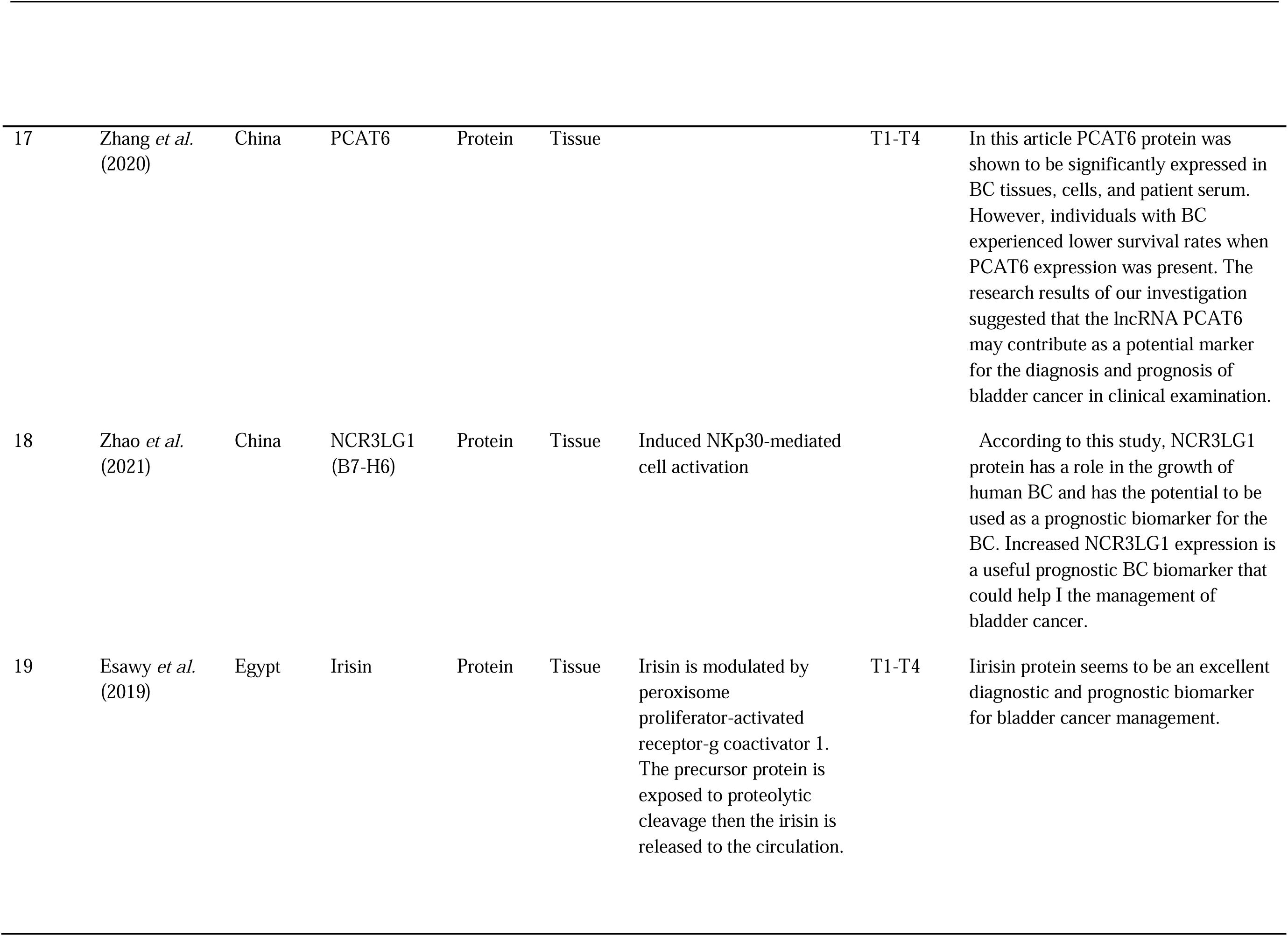
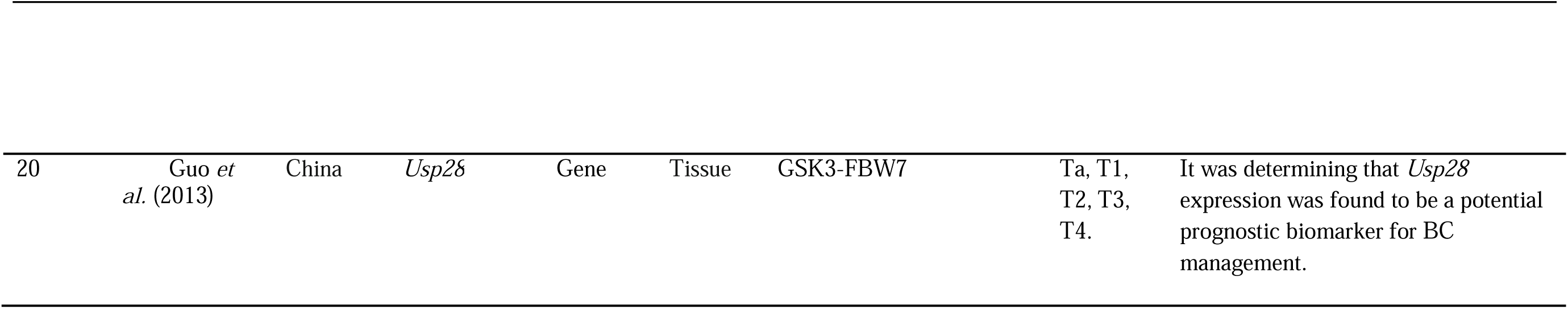
Summarized the data extracted and analysed from all the twenty selected studies. All studies were published from January 2012 onwards. Eleven articles focused on prognostic biomarkers, while nine articles focused on diagnostic biomarkers.

The studies collectively identified thirteen gene product biomarkers and seven protein product biomarkers. Gene biomarkers discovered included TEAD4, TPM1, TPM2, SKA3, ENO1, HYAL3, MTDH, EPDR1, hTERT, KRT7, SW, ARHGAP9, NXPH4, OTX1, BUB1, and Usp28. Protein biomarkers identified were A1AT, APOE, AG, CA9, IL8, MMP9, MMP10, PAI1, SDCI1, SDC1, VEGFA, CD73, TIP2, CXCL5, PCAT6, and NCR3LG1 (B7-H6).

These biomarkers were detected in various sample types, including tissue, urine, and blood, reflecting the diverse methodologies employed. The biomarkers are associated with multiple pathways such as the PI3K/AKT pathway, immune-related pathways, cell cycle pathways, and glycolysis pathways. These pathways play crucial roles in tumor development, immune cell infiltration, and cancer cell motility, which are critical for understanding the mechanisms underlying bladder cancer progression. The biomarkers were linked to different stages of bladder cancer, ranging from non-muscle invasive bladder cancer (NMIBC) to muscle-invasive bladder cancer (MIBC). For instance, TEAD4 was identified as a prognostic biomarker promoting epithelial to mesenchymal transition (EMT) in bladder cancer cells by activating the PI3K/AKT pathway, enhancing cell migration and invasion. TPM1 and TPM2 were highlighted as useful diagnostic indicators linked to immune cell invasion in bladder cancer, while SKA3 was noted for its association with immune infiltration affecting patient prognosis.

The studies also reported the potential of certain protein biomarkers for clinical application. A1AT, APOE, AG, CA9, IL8, MMP9, MMP10, PAI1, SDCI1, SDC1, and VEGFA were part of a multiplex Oncuria test that accurately identifies and monitors bladder cancer through urine samples. Similarly, CD73, an adenosine-producing cell surface enzyme found in blood, was proposed as a prognostic biomarker for bladder cancer.

Despite the promising findings, the implementation of these biomarkers in clinical practice is still under research. The studies collectively highlight the need for further investigation to validate the diagnostic and prognostic potential of these biomarkers and determine their clinical applicability.

### 4.3 Quality Assessment

The quality of the included studies was evaluated using the Quality Assessment of Diagnostic Accuracy Studies (QUADAS-2) tool. This tool assesses the risk of bias across four domains: patient selection, index test, reference standard, and flow and timing. A study was declared to have a low risk of bias in a domain if most of the review questions for that domain were answerable and scored positively. All twenty studies included in this systematic review were assessed for quality, and the majority were found to be of high quality. Specifically, eighteen of the included studies (90%) were rated as having a low risk of bias across all assessed domains. These studies provided clear and comprehensive information on patient selection, the index test, reference standards, and the flow and timing of the diagnostic process, thereby ensuring robust and reliable results.

Two studies (10%) were rated as having some concerns regarding bias, primarily due to incomplete reporting in the flow and timing domain. These concerns, however, were not significant enough to exclude the studies from the review. Instead, they indicate areas where future research can improve reporting standards to ensure complete transparency and reproducibility.

Overall, all twenty studies were deemed to have moderate to high quality, with the majority exhibiting low risk of bias. Consequently, they were included in the quantitative synthesis, providing a reliable basis for the systematic review’s findings on diagnostic and prognostic biomarkers for bladder cancer.

## Discussion

This systematic review identified twenty potential biomarkers for the diagnosis and prognosis of bladder cancer, consisting of thirteen gene products (TEAD4, TPM1, TPM2, SKA3, ENO1, HYAL3, MTDH, EPDR1, hTERT, KRT7, SW, ARHGAP9, NXPH4, OTX1, BUB1, and Usp28) and seven protein products (A1AT, APOE, AG, CA9, IL8, MMP9, MMP10, PAI1, SDCI1, SDC1, VEGFA, CD73, TIP2, CXCL5, PCAT6, NCR3LG1 (B7-H6), and irisin).

These biomarkers were identified through various methodologies using tissue, urine, and blood samples, and are involved in diverse pathways, including the PI3K/AKT pathway, immune-related pathways, and cell cycle pathways.

The study by Chi *et al*., (2022) highlighted TEAD4 as a potential prognostic biomarker that promotes epithelial to mesenchymal transition (EMT) in bladder cancer cells, enhancing cell migration and invasion through the PI3K/AKT pathway. This finding aligns with previous research suggesting the role of the PI3K/AKT pathway in cancer progression. Similarly, Yan *et al*., (2022) identified TPM1 and TPM2 as useful diagnostic indicators linked to immune cell invasion in bladder cancer, supporting the notion that immune system interactions play a critical role in tumor development.

SKA3, identified by Wang et al., (2022), was linked to immune infiltration and was reported to affect a patient’s prognosis for bladder cancer. This is consistent with other studies that have shown the involvement of immune cells, such as M2 macrophages and Th2 cells, in cancer progression. Huang *et al*., (2022) found that ENO1 was substantially correlated with tumor aggressiveness and abnormality, being overexpressed in bladder cancer. This upregulation of ENO1 predicted worse clinical outcomes, suggesting its potential as a diagnostic biomarker, which corroborates findings from other cancers where ENO1 has been implicated.

Liu *et al*., (2022) determined that a shorter overall survival among bladder cancer patients may be predicted by a greater HYAL3 expression level. This biomarker was linked to a variety of invading immune cells, further emphasizing the role of the immune environment in cancer prognosis. Zhou *et al*., (2012) suggested the possibility of using MTDH as a prognostic biomarker for bladder cancer patients, a finding that resonates with other studies on cancer progression markers. Hirasawa et al., (2021) demonstrated the diagnostic potential of a panel of urine-based proteins, including A1AT, APOE, AG, CA9, IL8, MMP9, MMP10, PAI1, SDCI1, SDC1, and VEGFA, which could be useful in bladder cancer management. This aligns with the trend towards non-invasive diagnostic techniques.

Yahyazadeh *et al*., (2021) reported the diagnostic potential of monitoring bladder cancer by evaluating the amounts of hTERT, SW, and KRT7 mRNA in urine. This method shows higher sensitivity than cytology, providing a non-invasive approach for diagnosing and following up on low-grade transitional cell carcinoma (TCC) of the bladder. Koivisto *et al*., (2019) identified CD73, an adenosine-producing cell surface enzyme, as a potential prognostic biomarker, supporting findings from other studies on adenosine’s role in cancer progression. Piao *et al*., (2020) identified ARHGAP9 as potential oncogenes in bladder cancer that may be used as unique prognostic markers, adding to the body of evidence on genetic markers in cancer classification.

Sun *et al*., (2022) discovered NXPH4 as a unique prognostic molecular marker linked to immune cell infiltration in bladder cancer. This finding could guide future research on bladder cancer tumor immunity. Jiang *et al*., (2022) determined that TIPE2 was overexpressed in clinicopathological diagnosis and prognosis in bladder urothelial cancer, suggesting its employment as a potential protein biomarker for disease management. Jiang *et al*., (2021) also identified OTX1 as a potential gene for the diagnosis and prognosis of bladder cancer, with carcinogenic effects through cell cycle-related pathways. Piao *et al*., (2021) proposed that BUB1 controls the G2/M transition to increase bladder cancer cell proliferation, potentially acting as a prognostic biomarker for NMIBC. Zhang *et al*., (2020) identified CXCL5 protein as a potential prognostic biomarker for bladder cancer tissue and a novel marker for diagnosis in urine.

Zhang *et al*., (2020) demonstrated that PCAT6 protein was significantly expressed in bladder cancer tissues, cells, and patient serum, with lower survival rates associated with its expression. This investigation suggested that lncRNA PCAT6 may serve as a potential marker for diagnosis and prognosis in clinical examinations. According to Esawy & Abdel-Samd, (2020), NCR3LG1 protein has a role in the growth of human bladder cancer and could be used as a prognostic biomarker, with increased expression being a useful indicator.

Lastly, Guo *et al*., (2014) determined that Usp28 expression was found to be a potential prognostic biomarker for bladder cancer. Despite the identification of several potential biomarkers in blood, tissue, and urine, their implementation in clinical application remains limited. Further studies are required to validate these biomarkers and establish their clinical utility.

## Conclusion

This systematic review identified twenty potential biomarkers that could aid in the detection and management of bladder cancer. Among these, thirteen were gene product biomarkers (TEAD4, TPM1, TPM2, SKA3, ENO1, HYAL3, MTDH, EPDR1, hTERT, KRT7, SW,

ARHGAP9, NXPH4, OTX1, BUB1, and Usp28) and seven were protein product biomarkers (A1AT, APOE, AG, CA9, IL8, MMP9, MMP10, PAI1, SDCI1, SDC1, VEGFA, CD73, TIP2, CXCL5, PCAT6, NCR3LG1 (B7-H6), and irisin). These biomarkers were identified in patients with bladder cancer and are involved in various pathways, highlighting their potential role in disease progression and prognosis.

The findings underscore the significant promise of these biomarkers in improving bladder cancer diagnosis and reducing the need for invasive procedures like cystoscopy. However, the studies reviewed provided limited details on the clinical application and validation of these biomarkers. Therefore, further research is essential to confirm their diagnostic and prognostic value, establish standardized protocols for their use, and integrate them into clinical practice.

In conclusion, while this review highlights the potential of gene and protein biomarkers in bladder cancer management, extensive validation and clinical trials are necessary to translate these findings into practical diagnostic tools and therapeutic targets. Future studies should focus on large-scale validation, the development of non-invasive testing methods, and the exploration of these biomarkers’ roles in personalized medicine to enhance patient outcomes in bladder cancer.

## Data Availability

All data produced in the present work are contained in the manuscript

## List of abbreviations

(ATP): Adenosine triphosphate
(BC): Bladder cancer
(BTA): Bladder tumour antigen
(CCD1): Cyclin D1
(EMT): Epithelial to mesenchymal transition
(IVU): Intrasonography intravascular urography
(MIBC): Muscles invasive bladder cancer
(NMIBC): Non-muscle invasive bladder cancer
(MP22): Nuclear matrix protein 22

## Declaration

### Authors’ contributions

Conceived and designed the study: UA. Performed literature mining and wrote the first draft: UM and UA. Reviewed and revised the draft: BI. Conducted the analysis: UM. Reviewed the final draft: AAA and HUL.

